# The role of Rwanda Field Epidemiology and Laboratory Training Program graduates and residents in response to the first Marburg Virus Disease outbreak in Rwanda

**DOI:** 10.1101/2025.03.11.25323792

**Authors:** Edouard Ruseesa, Aphrodis Hagabimana, Fredrick Ntirenganya, Peace Kinani, Ruton Hinda, Emmanuel Nshimiyimana, Claude Niyoyita, Angela Umutoni, Caroline Stamatakis, Hugues Valois Mucunguzi, Edson Rwagasore

## Abstract

**Background:** Rwanda Field epidemiology and laboratory training programs (RFELTP) build capacity to improve disease surveillance, respond to public health emergencies, and inform evidence-based decision-making to mitigate the impact of epidemics and other public health events. On September 27, 2024, Rwanda declared its first Marburg virus disease (MVD) outbreak. MVD disease is a severe haemorrhagic fever with a fatality rate ranging from 24% to 90%. We describe the role played by the RFELTP in the outbreak response.

**Methods:** In November 2024, we provided a self-administered questionnaire to all residents and graduates of RFELTP. The questionnaire included sections on sociodemographic, experience in outbreak response, and MVD outbreak response experiences. Descriptive statistical analysis was performed, and the data were analysed using Stata version 16.

**Results:** Among the 523 total participants, 34.8% supported MVD response activities. Residents and graduates supported the response in all 30 districts of Rwanda, mainly at the national and sector levels. Most supported the epidemiology and surveillance 41.2%, data management 19.2%, infection prevention control 14.3%, and leadership and coordination pillars 10.4%. Nearly all 96.7% felt adequately prepared by their RFELTP to handle outbreak responses.

**Conclusion:** The RFELTP graduates and residents proved to be a critical resource in the MVD outbreak response, demonstrating the capacity to respond to public health emergencies. Continued investment in this training program is essential for enhancing global health security and strengthening capabilities to respond to public health emergencies.

## Background

The RFELTP is a competency-based training program aimed at strengthening field epidemiology capacity at all levels of the health system to prevent, detect, investigate and respond to public health priority issues [1]. RFELTP is led by the Government of Rwanda, with assistance from the University of Rwanda’s School of Public Health, African Field Epidemiology Network, as well as technical support and funding from the U.S. Centers for Disease Control and Prevention. The program consists of three tiers: Frontline, Intermediate, and Advanced. Established in 2009, the Advanced Tier consists of both didactic components and field placements that facilitate competency development in outbreak investigation, surveillance systems, data analysis, protocol-based studies, and scientific communication. To date, the program has trained 91 field epidemiologists who are primarily deployed at the national and district levels to provide public health leadership and technical expertise. The Intermediate Tier was launched in 2022 as a nine-month training program that has a similar curriculum structure as the Advanced training, but mainly targets district-level health officials. As of December 2024, 33 Intermediate RFELTP residents have been trained. The Frontline Tier was introduced in 2020 as a three-month program, largely in response to the COVID-19 pandemic. It focuses on sub-national levels of the health sector and consists of three on-site training workshops, and has two field placements during which participants complete field projects in their primary work sites throughout Rwanda. Through the Frontline Tier, 399 participants have been trained, including clinical directors from hospitals, integrated disease surveillance and response officers, monitoring and evaluation officers, health promotion officers, animal resources officers and others. In total, the RFELTP has trained 523 professionals (including graduates and current residents) in field epidemiology across the three tiers (Frontline: 399, Intermediate: 33, Advanced: 91). Graduates serve at various health administrative levels in Rwanda and make up the country’s primary roster of surge staff available to respond to urgent public health situations.

MVD is a severe and often fatal haemorrhagic fever caused by infection with the Marburg virus [2-3]. MVD, which belongs to the same family (*Filoviridae*) as Ebola virus, has a case-fatality rate that ranges from 24% to 90% [4-6]. Egyptian fruit bats (*Rousettus aegyptiacus*) are the reservoirs of MV [7-9]. Transmission of the virus occurs through either direct or indirect contact between fruit bats and humans and through human-to-human transmission with an estimated 2–21 days incubation period [10].

On 27 September 2024, Rwanda’s Ministry of Health (MoH) declared the nation’s first-ever MVD outbreak. In total, 66 cases were confirmed, including 15 deaths, resulting in a case-fatality rate of 22.7% [11]. All confirmed cases were detected from the three districts in the capital, Kigali City. Kigali is characterized by its dense population and significant influx of people from various districts and foreign countries [12], creating conditions that potentially increase the risk of disease transmission. Health workers from two health facilities in Kigali, King Faisal Hospital and the University Teaching Hospital of Kigali, accounted for the majority of confirmed cases. Confirmed cases who were not healthcare workers were also linked to these health facilities [13].

Effectively managing an MVD outbreak requires a comprehensive strategy encompassing several key measures. These include early case identification, patient isolation and treatment, disease surveillance, systematic contact tracing and quarantine, infection prevention and control, robust laboratory diagnostic capabilities, safe and dignified burials, and effective risk communication and community engagement [14]. The effectiveness of these interventions depends on a well-trained public health workforce [15]. The World Health Organisation (WHO) also recognizes a strong public health workforce as critical to compliance with the 2005 International Health Regulations [16].

We aimed to describe the role of RFELTP graduates and residents in response to the MVD outbreak in Rwanda from September to mid-November 2024.

## Methodology

We described the roles of RFELTP residents in the MVD response to illustrate their impact on emergency response. A complete list of all 523 graduates and residents from the Advanced, Intermediate, and Frontline RFELTP tiers was obtained from the University of Rwanda and Rwanda Biomedical Centre (RBC), which oversees the RFELTP program.

From 1^st^ to 30^th^ November 2024, we conducted an online survey using a questionnaire designed through the Kobo Toolbox platform [16]. All 523 graduates and residents (individuals still in the RFELTP training) were contacted through phone calls and emails to participate in the online survey. The research team provided guidance and support by phone to answer questions participants raised, to ensure complete and accurate responses.

The survey questionnaire consisted of two main parts. The first section collected demographic data (age, sex, RFELTP training level, training cohort, professional background, current employment, and years of experience in epidemiology and outbreak response). The second section focused on the participants’ specific roles, activities, and challenges encountered during the MVD outbreak response. The survey collected information on involvement across the different response pillars, including leadership and coordination, epidemiological surveillance, laboratory, case management, infection prevention and control, risk communication, and data management.

Descriptive statistics (frequencies and percentages) were calculated to summarize the participants’ sociodemographic characteristics and their involvement in various aspects of the MVD outbreak response. Data were analysed using Stata version 16 [17] and presented in tables, a map and graphs.

### Ethics approval and consent to participate

The information reported in this article represents data collected during the MVD outbreak response. An outbreak investigation is regarded as an emergency activity and was endorsed by MOH. Permission to analyse and publish this information was sought and granted by Rwanda

Biomedical Centre (RBC). In accordance with the Declaration of Helsinki, participation in the survey was voluntary, and informed consent was obtained from all participants prior to completing the survey. To maintain the confidentiality of respondents, strict measures were implemented to safeguard participant data. During the data collection phase, all personal identifying information was anonymized.

## Results

All of the 523 RFELTP graduates and residents were emailed the survey. Among 209 (40.0%) RFELTP graduates and residents who responded to the survey, a total of 182 (87.1%) were involved in the MVD response that included; 28.6% the advanced tier, 14.8% intermediate tier and 56.6%. Frontline Tier. The majority 55.5% were aged between 35-44 years, 65.9% were male, and 57.3% had more than five years of experience in public health or epidemiology (Table 1).

**Table 1.** Sociodemographic characteristics of the participants, Rwanda, 2024.

| Variable | Category | Frequency (n) | Percentage (%) |
| --- | --- | --- | --- |
| Age group | 25-34 | 44 | 24.2 |
|  | 35-44 | 101 | 55.5 |
|  | 45-54 | 32 | 17.6 |
|  | ≥55 | 5 | 2.7 |
| Sex | Female | 62 | 34.1 |
|  | Male | 120 | 65.9 |
| Highest tier of FELTP | Advanced | 52 | 28.6 |
|  | Intermediate | 27 | 14.8 |
|  | Frontline | 103 | 56.6 |
| Training status | Graduate | 118 | 64.8 |
|  | Residents | 64 | 35.2 |
| Professional background | Allied Health Sciences | 81 | 44.5 |
|  | Animal Health/Veterinary | 8 | 4.4 |
|  | Data Science | 28 | 15.4 |
|  | Medical doctor | 20 | 11.0 |
|  | Nurse | 45 | 24.7 |
| **Current level of employment** | National | 53 | 29.4 |
|  | Province | 8 | 4.4 |
|  | District | 88 | 48.9 |
|  | Sector | 31 | 17.3 |
| **Type of employment** | Civil servant | 162 | 89.0 |
|  | Partners | 14 | 7.7 |
|  | Academia | 4 | 2.2 |
|  | Private | 2 | 1.1 |
| **Years of experience in public health/epidemiology** | > 5 | 105 | 57.7 |
|  | 3-5 | 37 | 20.3 |
|  | 1-3 | 32 | 17.6 |
|  | <1 | 8 | 4.4 |

### Role of Rwanda Field Epidemiology and Laboratory Training Program graduates and residents in Marburg virus disease response, Rwanda, 2024

An initial group of four RFELTP graduates were deployed as part of a team to conduct a preliminary case investigation on 25 September 2024. The outbreak was declared by the MoH on 27 September 2024. On the same day, ten additional RFELTP graduates, primarily those who worked at the national level (MoH and RBC), and 5 residents were deployed to support the National MVD command post in case investigation and contact tracing. On 2 October 2024, additional graduates and residents from sub-national and private institutions were mobilized to provide support at the national level within the MVD Command Post. All RFELTP advanced tier graduates and residents were first contacted to determine availability. For the residents and graduates who confirmed availability, formal invitation emails were sent to their respective institutions, and all were able to support the response. The RFELTP graduates and residents from the intermediate and frontline tiers who participated in the MVD response supported preparedness and response activities at the health sector levels where they were working (Fig 1.).

**Fig 1.**
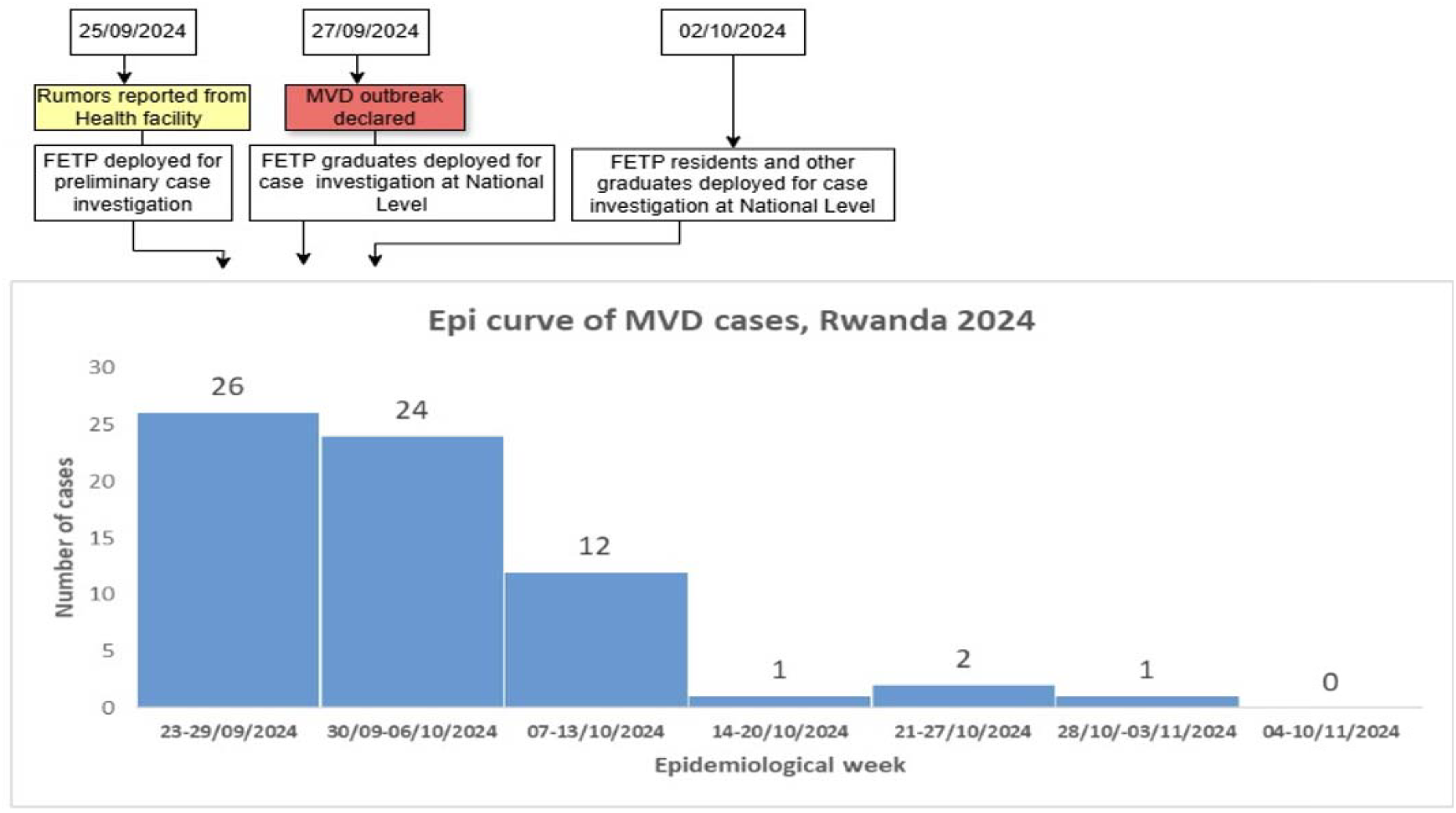
Timeline of Rwanda Field Epidemiology and Laboratory Training Program graduates and resident’s deployment during the Marburg virus disease outbreak in Rwanda, September-November 2024.

The RFELTP team supported MVD response in all 30 districts of Rwanda; however, the majority were deployed in Kigali City which was the epicenter of the outbreak (Fig 2.).

**Fig 2.**
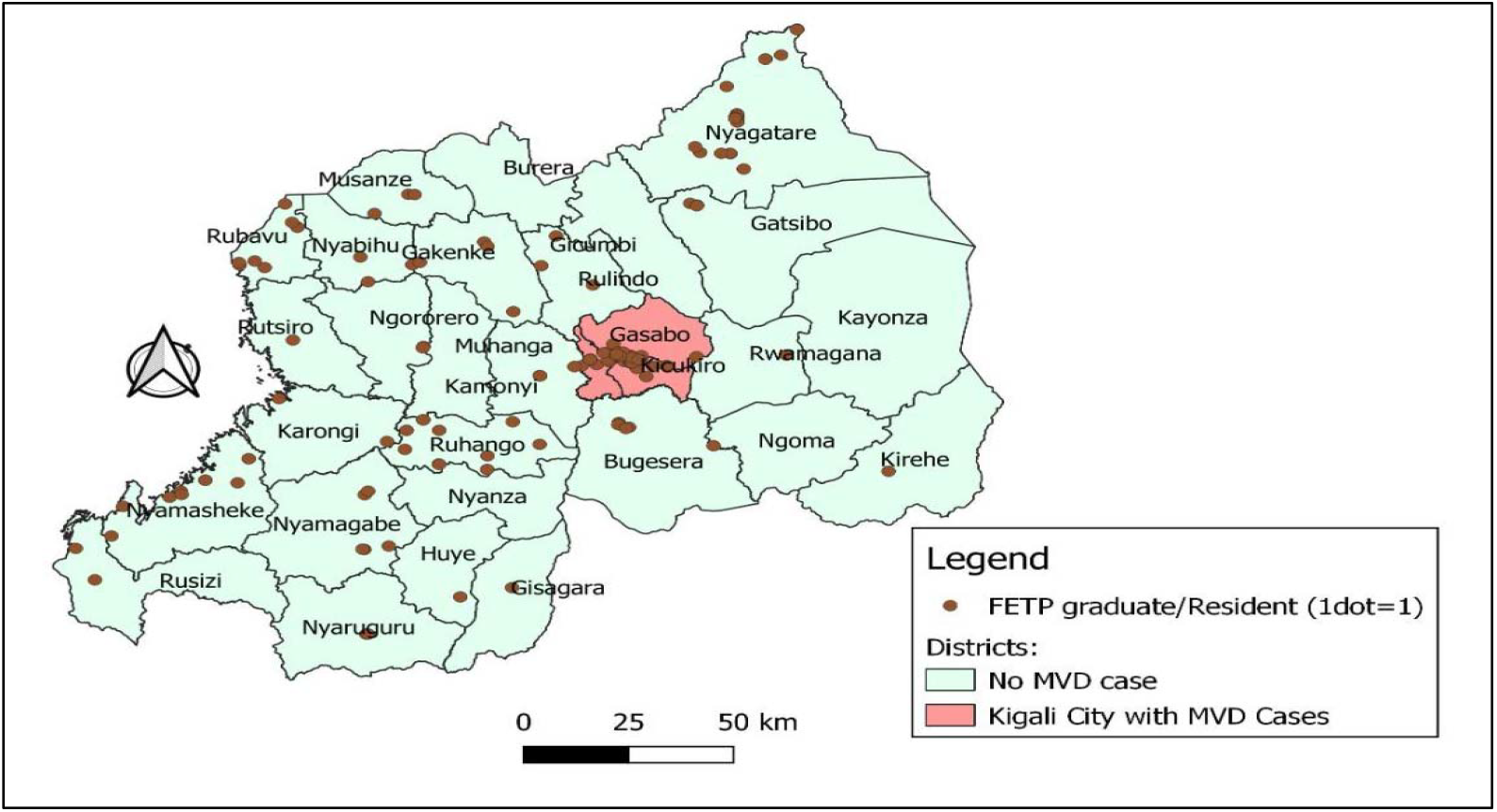
Distribution of Rwanda Field Epidemiology and Laboratory Training Program graduates and residents in Marburg virus disease response (n=182), by district supported, 2024

The participants involved in the MVD response were from all RFELTP tiers: Advanced, Intermediate, and Frontline. The majority of RFELTP Advanced graduates and residents supported response operations at the national level, while the RFELTP Intermediate and Frontline mainly provided support at the district and sector levels (Fig 3.).

**Figure 3:**
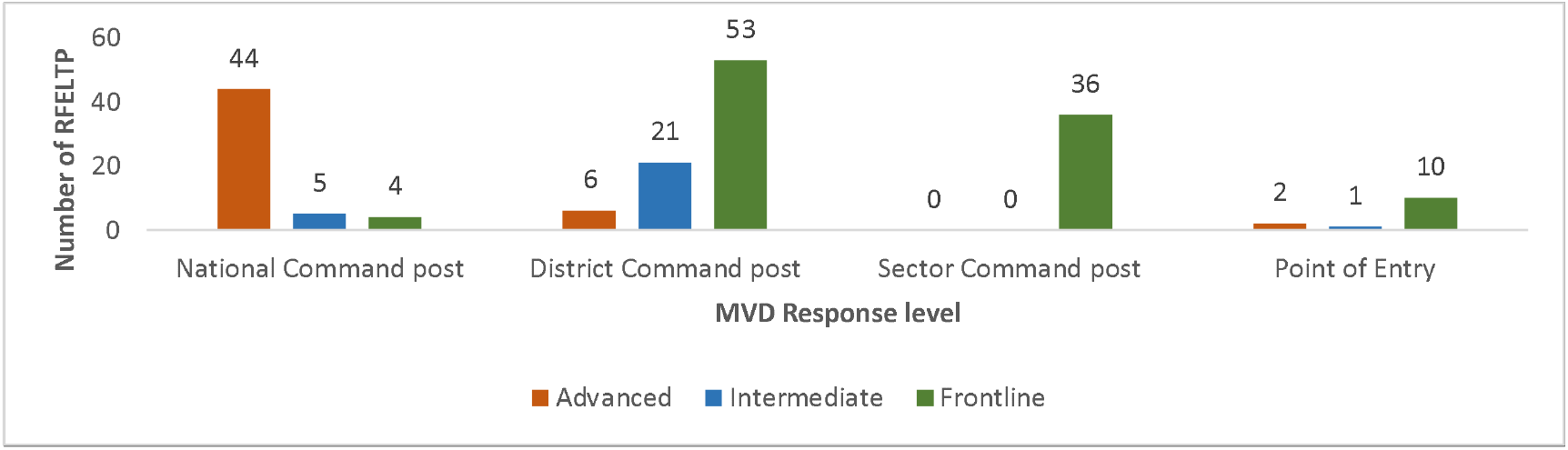
Distribution of Rwanda Field Epidemiology and Laboratory Training Program Tiers by Marburg virus disease response levels, Rwanda, 2024.

RFELTP graduates and residents were primarily involved in epidemiological surveillance (n=75, 41.2%), data management (n=35, 19.2%), infection prevention and control (n=26, 14.3%) and leadership and coordination (n=19, 10.4%) (Table 2).

**Table 2:** Rwanda Field Epidemiology and Laboratory Training Program participation in Marburg virus disease response Pillars, September to November 2024, Rwanda.

| **Pillar** | **Frequency**<br>**(n)** | **Percentage**<br>**(%)** |
| --- | --- | --- |
| Epidemiological surveillance | 75 | 41.2 |
| Data management | 35 | 19.2 |
| Infection prevention and control | 26 | 14.3 |
| Leadership and coordination | 19 | 10.4 |
| Laboratory | 13 | 7.1 |
| Risk communication and community engagement | 8 | 4.4 |
| Case management | 3 | 1.7 |
| Vaccination | 3 | 1.7 |

RFELTP graduates and residents provided support to District Emergency Operation Centres throughout Rwanda to deliver MVD trainings (Figure 4.). RFELTP graduates and residents visited schools to train teachers, students, and community health workers on MVD signs and symptoms, transmission, preventive measures, and community active case search. Healthcare providers were trained on case investigation forms, donning and doffing of personal protective equipment, hand washing practices, decontamination, waste segregation, final disposal, and other safety precautions. Youth volunteers, a group of young people who contribute to Rwanda’s development without seeking compensation, and other groups of people such as District Administration Security Support Organ staff and the Rwanda National Police staff at the local level were trained to provide support in community sensitization of MVD.

**Fig 4:**
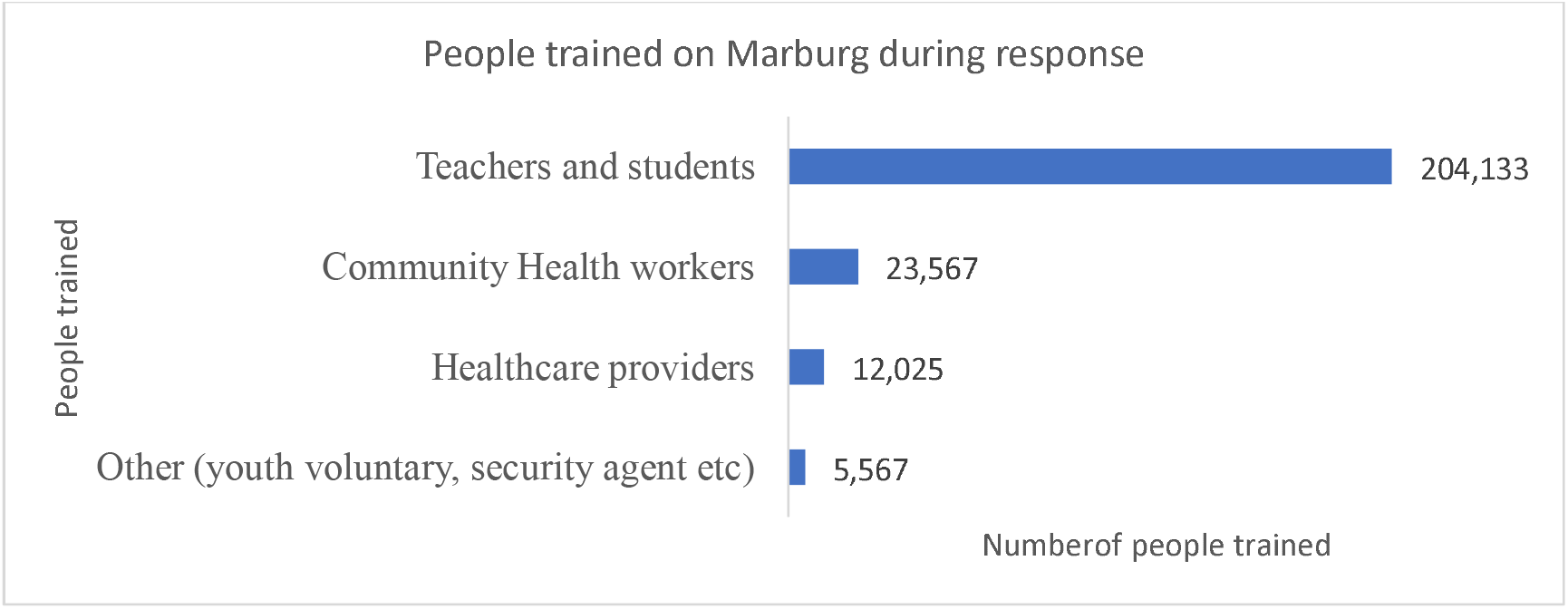
Number of people trained by Rwanda Field Epidemiology and Laboratory Training Program Team on Marburg virus disease preparedness and response, Rwanda, 2024

The majority 96.7% of RFELTP graduates and residents felt adequately prepared by their RFELTP training to handle outbreak response. The greatest strengths of their training identified by the REFLTP graduates were in the areas of case investigation, data collection and analysis, and contact tracing. Participants identified advanced analytical data analysis as the most recommended area for further training improvements to improve the role of RFELTP graduates and trainees in future outbreak responses (Table 3).

**Table 3.** Preparedness, Strengths, and Area for improvement by the Rwanda Field Epidemiology and Laboratory Training Program participants, Rwanda, 2024.

| Variable | Category | Frequency (n) | Percentage (%) |
| --- | --- | --- | --- |
| Feel adequately prepared by REFLTP training to handle the response | Yes | 176 | 96.7 |
|  | No | 6 | 3.3 |
| Which aspects of your REFLTP training helped you perform your role during the MVD outbreak response? | Case Investigation | 137 | 20.9 |
|  | Data Collection and Analysis | 130 | 19.8 |
|  | Contact tracing methods | 109 | 16.6 |
|  | Field Safety Protocols | 98 | 14.9 |
|  | Coordination and Leadership | 96 | 14.6 |
|  | Risk Communication Techniques | 87 | 13.2 |
| What do you think is the biggest strength of REFLTP graduates/trainees in outbreak response? | Case investigation and contact tracing | 92 | 33.4 |
|  | Surveillance analysis | 74 | 26.7 |
|  | Communication and community engagement skills | 54 | 19.6 |
|  | Epidemiological skill | 48 | 17.4 |
|  | Laboratory skills | 8 | 2.9 |
| Areas would you recommend for further REFLTP training improvements? | Advanced Data Analysis Skills | 93 | 24.4 |
|  | Risk Communication in Outbreak Settings | 74 | 19.4 |
|  | Mental Health and Resilience Training | 68 | 17.8 |
|  | Rapid Response Field Protocols | 62 | 16.2 |
|  | Community Engagement Techniques | 52 | 13.6 |
|  | Inter-Agency coordination | 33 | 8.6 |

## Discussion

This study describes the role of RFELTP graduates and residents during the 2024 Rwanda MVD outbreak. The deployment and response support of all trained epidemiologists, representing diverse professional backgrounds and geographic locations, underscores the importance of having a well-trained public health workforce. Without RFELTP, this considerable group of trained individuals would not have been available to enhance Rwanda’s capacity to respond effectively to this public health emergency.

Survey respondents identified key strengths of RFELTP graduates and residents, such as case investigation, contact tracing, surveillance analysis, and community engagement. These competencies were instrumental in the effective management of the MVD outbreak, particularly in areas such as surveillance and case investigation, where RFELTP teams established a strong reporting mechanism, successfully identified all contacts, tracked the index case, and mapped high-risk groups [19]. These areas of engagement align with the core competencies of the RFELTP, which emphasize data-driven outbreak investigation and management [1,15,18] and are similar to engagement in previous success stories that other Field Epidemiology Training Programs (FETPs) worldwide have demonstrated in fulfilling similar roles during significant outbreaks. For example, FETPs in Liberia proved effective in managing Ebola virus outbreaks through robust case investigation and contact tracing initiatives [20] and in the 2023 MVD outbreak in Tanzania by providing support in surveillance and contact tracing [21].

While most of the advanced tier graduates and residents provided support in Kigali City where all confirmed MVD cases were detected [13], other graduates and residents provided response support in all 30 districts of the country, including at their respective workplaces. This geographical distribution of RFELTP graduates and residents enabled localized interventions that were tailored to the specific needs of communities, including the training of more than 245,000 individuals across the country. The RFELTP team’s role in door-to-door screening and community education aimed to combat inaccuracies in circulating information and build community trust in managing transmission risks. These efforts highlight the benefits of having a three-tiered program that trains individuals at all levels of the health sector to provide support not only at the national level, but in the field and on the ground for enhanced implementation of interventions.

The participants reported high confidence in their RFELTP training to handle the outbreak response with many assuming leadership and coordination roles. RFELTP residents and graduates served as national leads of the surveillance pillar, partner coordination pillar, case investigation team, and contact follow-up team. At the district level, they served as case investigation, infection prevention and control, and laboratory team leads. This high level of reported confidence in their training preparedness and their representation in the response in leadership positions suggests that the program is achieving one of its primary objectives, to effectively build capacity to respond to public health emergencies [1].

Despite the demonstrated strengths of the RFELTP program, participants identified areas for future improvement in the RFELTP curriculum, including advanced data analysis skills, enhanced risk communication in outbreak settings, and mental health and resilience training. These suggestions indicate opportunities for improving the RFELTP curriculum to further strengthen outbreak response capabilities.

This study has several limitations. The use of self-reported data and the provision of phone guidance by the research team may introduce the possibility of response bias and limit the generalizability of the findings. However, the phone support was solely intended to ensure participants fully understood the questions, in order for them to complete the survey. Additionally, the study was conducted within a short time frame of three months, which may not capture the full complexity of the outbreak response.

## Conclusion

Our findings emphasize the essential role of structured field epidemiology training programs like RFELTP in coordinated responses to emerging infectious disease threats. The multidisciplinary approach and practical skills demonstrated by RFELTP graduates and residents provide a model for building resilient public health systems. The RFELTP has proven to be a valuable resource in outbreak response, producing well-trained, adaptable professionals capable of managing complex public health emergencies. Continued investment and iterative improvements in the RFELTP will not only enhance Rwanda’s public health workforce capacity but will also serve as a model for similar programs in other countries.

## Data Availability

All data generated or analysed during this study are included in this article

## Consent for publication

Not applicable

## Availability of data materials

All data generated or analysed during this study are included in this article

## Author contributions

E. R, A .H, F.N, E.N, C.N designed the study and conducted the literature review and helped to prepare the introduction and methods sections of the text. E. R, A .H, F.N, E.N, C.N, P.K, R.H, H.V.M, E.R directed the study’s implementation. E. R, A .H, F.N, E.N, C.N, P.K, R.H, A.U, C.S, E.R designed the analytical strategy and helped to interpret the findings. C.N, P.K, R.H, A.U, C.S, E.R drafting the original draft and editing.

## Acknowledgments

Authors are thankful to the Rwanda FELTP participants, who voluntarily accepted to complete the questionnaires. The authors are also grateful to the MoH, RBC for having granted the permission to conduct this study. Our thanks also extend to US CDC, African Field Epidemiology Network and the University of Rwanda.

## Abbreviation

MVD: Marburg virus disease
MoH: Ministry of Health
RFELTP: Rwanda Field epidemiology and laboratory training programs
RBC: Rwanda Biomedical Centre
WHO: World Health Organisation

